# Strategies to Reach Employees Nurturing Guidance and Trust in Healthcare (STRENGTH): Understanding Employee Drivers for Employer-Based Influenza Vaccination Programs

**DOI:** 10.64898/2026.07.13.26357995

**Authors:** Arthur Sanchez, Kent Isakari, Morgan Keefe, Marcia Isakari, Amy M. Sitapati

## Abstract

**Background:** Vaccine hesitancy and logistical barriers continue to limit influenza vaccination uptake among healthcare personnel. The STRENGTH initiative evaluated employee drivers of influenza vaccine acceptance in healthcare workgroups with lower vaccination compliance.

**Methods:** This mixed-methods program evaluation used a custom Epic electronic medical record dashboard to identify workgroups with lower influenza vaccination rates, followed by focus groups, targeted worksite vaccination clinics, and an anonymous survey. Survey data from 73 respondents were analyzed using descriptive statistics, Mann-Whitney U tests, Kruskal-Wallis tests, chi-square or Fisher exact tests, and Spearman rank correlations.

**Results:** Respondents had a mean age of 44.8 years (SD 11.3); 36 of 70 respondents reporting binary gender were male. Convenience of worksite vaccination received the highest importance ratings (mean 4.72/5), followed by protection from influenza (mean 4.38/5). Both were rated significantly above the neutral midpoint of 3 (Wilcoxon p<0.001). The strongest observed Spearman correlations were between peer pressure and leadership participation (rho=0.47) and between protection from influenza and willingness to receive a future combined influenza/COVID-19 vaccine (rho=0.42).

**Conclusions:** Convenience and perceived protection were the most important vaccination drivers. Employer-sponsored workplace vaccination programs can reduce logistical barriers while providing opportunities for targeted education and trust-building.

## Introduction

Influenza remains a significant occupational and public health concern in healthcare settings. Influenza vaccination has been found to be the most effective preventive strategy, reducing laboratory confirmed influenza by 64% and absenteeism rate by 37% among healthcare workers [1]. Healthcare personnel have increased opportunities for exposure to respiratory pathogens and may transmit influenza to medically vulnerable patients. Annual vaccination is therefore an important preventive strategy for both employee health and patient safety.

Despite longstanding recommendations supporting influenza vaccination, uptake among healthcare workers can vary across occupational groups and work settings. Vaccine acceptance is influenced by perceived risk, trust, concerns about adverse effects, social and organizational norms, and practical access barriers. Workplace vaccination programs are a promising strategy because they can reduce logistical barriers and deliver preventive services in settings where employees already spend substantial time [2, 3, 5].

The Strategies to Reach Employees Nurturing Guidance and Trust in Healthcare (STRENGTH) initiative was developed to better understand employee drivers of influenza vaccination and to inform employer-based interventions. The objectives were to identify perceived barriers to vaccination, characterize motivators of vaccine acceptance, evaluate attitudes toward future combined influenza/COVID-19 vaccination, and generate evidence for targeted occupational health programming.

## Methods

### Study design and setting

This was a mixed-methods quality improvement and program evaluation project conducted at UC San Diego Health. Phase I used a custom Epic electronic medical record dashboard to track real-time influenza vaccination rates by demographic and employee group characteristics. The dashboard enabled occupational health staff to identify workgroups with lower vaccination uptake in real time and prioritize targeted interventions during the campaign. Phase II focused on lower-compliance employee groups identified in Phase I and included focus groups, targeted on-site vaccination clinics, and anonymous surveys. In accordance with regulations at 45 CFR 46, the IRB determined that this project did not meet the DHHS definition of human subjects research under 45 CFR 46.

### Survey measures

Survey items included demographics, worksite and work setting, prior fall 2023 influenza vaccination status, and a 5-point importance rating for six vaccination factors: protection from influenza, peer pressure from coworkers, work leaders also getting vaccinated, convenience of getting vaccinated at the worksite, free incentives or rewards, and concern about side effects. Respondents were also asked whether they would receive a future combined influenza/COVID-19 vaccine.

### Statistical analysis

Categorical variables were summarized using counts and percentages; continuous variables were summarized using means and standard deviations. One-sample Wilcoxon signed-rank tests compared each Likert item with the neutral midpoint of 3. Mann-Whitney U tests compared Likert ratings between employees who did and did not report prior influenza vaccination. Kruskal-Wallis tests compared Likert ratings across future combination-vaccine response groups (yes, no, not sure). Chi-square tests or Fisher exact tests evaluated categorical associations. Spearman rank correlations assessed monotonic relationships among age, prior vaccination, future combination-vaccine willingness, and Likert ratings. Spearman correlations are reported with Benjamini-Hochberg false-discovery-rate (FDR) adjusted q-values as exploratory analyses.

## Results

### Participant characteristics

A total of 73 employees completed the survey. Age was reported by 71 respondents; mean age was 44.8 years (SD 11.3; median 45; range 23-66)

**Table 1.** Race distribution of survey respondents. Missing values are retained to show denominator transparency.

| Race | n | % |
| --- | --- | --- |
| White | 27 | 37.0% |
| Missing | 12 | 16.4% |
| Hispanic | 9 | 12.3% |
| Asian Pac Island | 8 | 11.0% |
| Other | 6 | 8.2% |
| Prefer not to say | 5 | 6.8% |
| Black or African American | 5 | 6.8% |
| American Indian/ Alaskan | 1 | 1.4% |

**Table 2.** Gender distribution of survey respondents.

| Gender | n | % |
| --- | --- | --- |
| Male | 36 | 49.3% |
| Female | 34 | 46.6% |
| Missing | 3 | 4.1% |

### Vaccination factors

**Table 3.** Likert-scale ratings of factors influencing influenza vaccination decisions. The “p vs 3” reports results from a one-sample Wilcoxon signed-rank test evaluating whether responses differed from the neutral midpoint of 3 on the five-point Likert scale. The “p by prior vaccination status” reports results from a Mann–Whitney U test comparing factor ratings between employees who reported receiving the 2023 influenza vaccine and those who reported not receiving it.

| Factor | n | Mean | SD | Median | IQR | Counts 1-5 | p vs 3 | p by prior vaccine status |
| --- | --- | --- | --- | --- | --- | --- | --- | --- |
| Protection from influenza | 71 | 4.38 | 1.08 | 5.0 | 4-5 | 2, 4, 9, 7, 49 | <0.001 | 0.550 |
| Peer pressure from coworkers | 70 | 1.90 | 1.36 | 1.0 | 1-3 | 45, 5, 10, 3, 7 | <0.001 | 0.771 |
| Work leaders also vaccinated | 69 | 2.90 | 1.61 | 3.0 | 1-5 | 23, 6, 14, 8, 18 | 0.501 | 0.993 |
| Convenience of worksite vaccination | 70 | 4.72 | 0.70 | 5.0 | 5-5 | 0, 2, 4, 6, 58 | <0.001 | 0.513 |
| Free incentives or rewards | 70 | 2.68 | 1.52 | 3.0 | 1-4 | 24, 10, 14, 9, 13 | 0.075 | 0.093 |
| Concern about side effects | 72 | 2.52 | 1.56 | 2.0 | 1-4 | 29, 11, 12, 6, 14 | 0.016 | 1.000 |

Likert scale ratings of factors: Convenience of workplace vaccination was the strongest driver of vaccination decisions. Protection from influenza was the second most important factor. Peer pressure from coworkers was the least important factor. Leadership participation, incentives, and concerns about side effects had intermediate importance.

p vs 3: Employees prioritized convenience and protection from influenza as important influences on vaccination decisions, while peer pressure was considered unimportant. Leadership participation and financial incentives were viewed approximately neutrally.

p by prior vaccine status: None of the six vaccination factors differed significantly between employees who reported receiving the 2023 influenza vaccine and those who did not.

**Figure 1.**
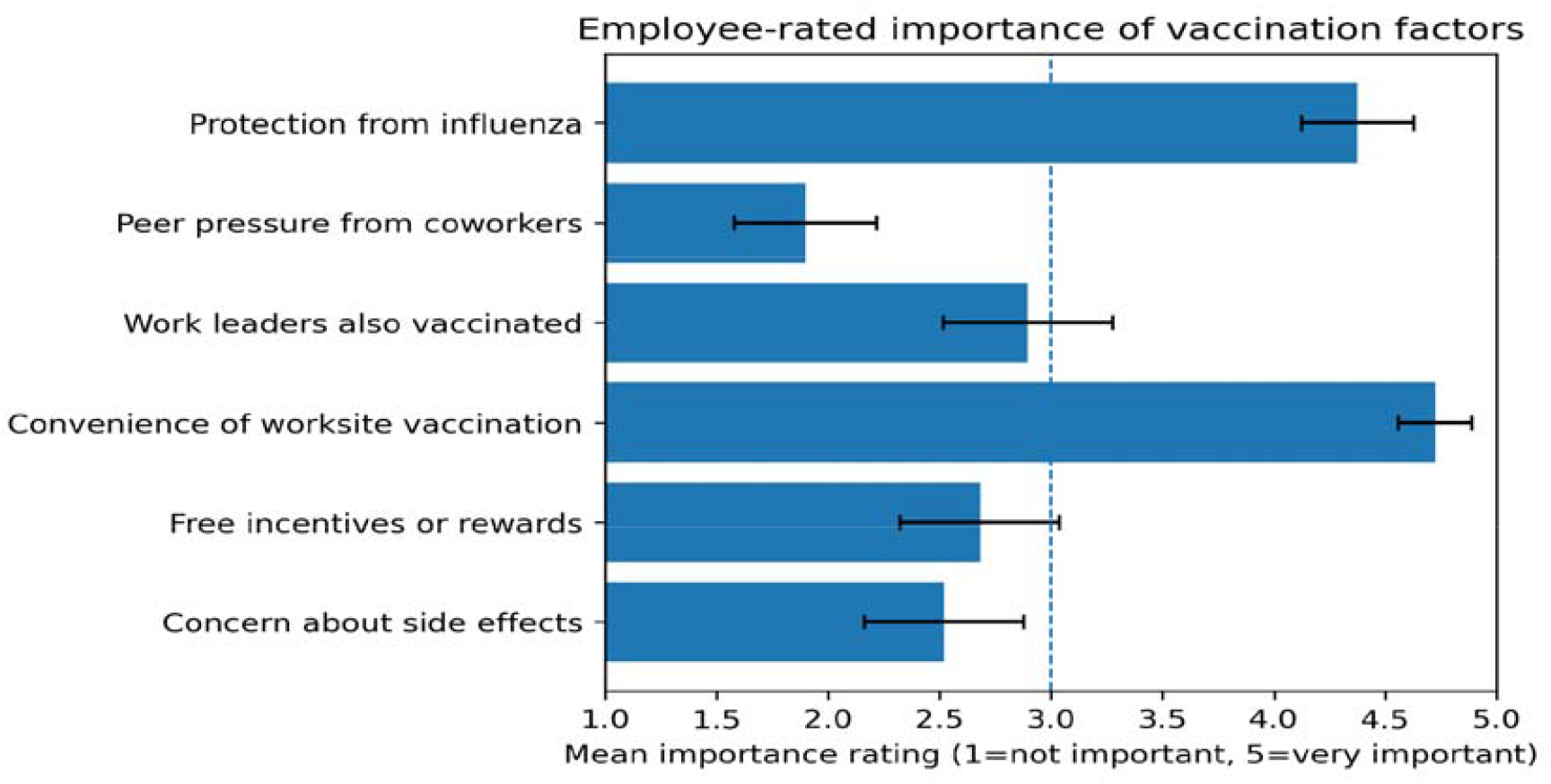
Mean importance ratings with 95% confidence intervals. The dashed reference line indicates the neutral midpoint of 3.

Convenience of worksite vaccination had the highest mean rating (4.72/5), with 58 of 70 respondents rating it as 5. Protection from influenza also scored highly (4.38/5). Peer pressure had the lowest mean rating (1.90/5).

### Future combined influenza/COVID-19 vaccine acceptance

**Table 4.** Respondent willingness to receive a future combined influenza/COVID-19 vaccine.

| Future combined vaccine response | n | % |
| --- | --- | --- |
| Yes | 36 | 49.3% |
| Not sure | 10 | 13.7% |
| No | 20 | 27.4% |
| Missing | 3 | 4.1% |

**Figure 2.**
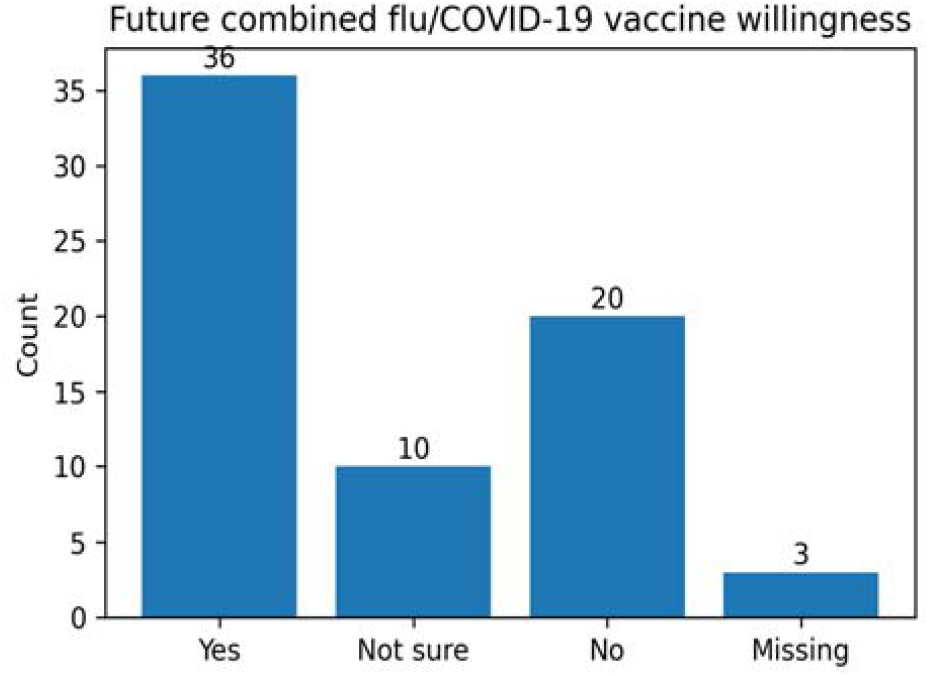
Distribution of future combination-vaccine willingness.

**Table 5.**
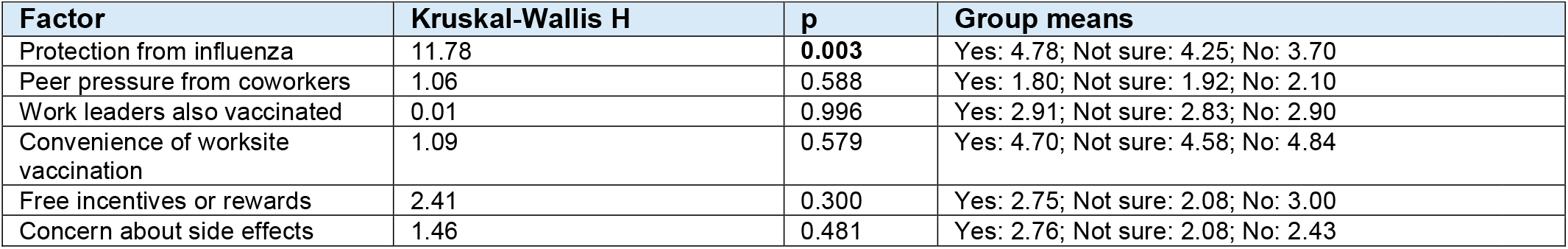
Differences in vaccination-factor ratings across future combination-vaccine willingness groups.

### Correlation and association analyses

**Table 6.** Strongest exploratory Spearman correlations by absolute rho. FDR q-values adjust for the set of tested correlations.

| Variable 1 | Variable 2 | n | Spearman rho | p | FDR q |
| --- | --- | --- | --- | --- | --- |
| Peer pressure from coworkers | Work leaders also vaccinated | 69 | 0.47 | **<0.001** | 0.002 |
| Combined flu/COVID vaccine willingness | Protection from influenza | 68 | 0.42 | **<0.001** | 0.007 |
| Work leaders also vaccinated | Concern about side effects | 69 | 0.34 | **0.004** | 0.036 |
| Peer pressure from coworkers | Concern about side effects | 70 | 0.34 | **0.004** | 0.036 |
| Age | Prior flu vaccination | 69 | 0.33 | **0.005** | 0.036 |
| Peer pressure from coworkers | Free incentives or rewards | 69 | 0.31 | **0.011** | 0.064 |
| Free incentives or rewards | Concern about side effects | 70 | 0.30 | **0.013** | 0.066 |
| Prior flu vaccination | Free incentives or rewards | 68 | -0.21 | 0.091 | 0.410 |
| Age | Work leaders also vaccinated | 68 | 0.18 | 0.133 | 0.534 |
| Age | Free incentives or rewards | 69 | -0.14 | 0.252 | 0.718 |
| Protection from influenza | Peer pressure from coworkers | 69 | -0.13 | 0.273 | 0.718 |
| Convenience of worksite vaccination | Concern about side effects | 70 | -0.13 | 0.283 | 0.718 |

**Figure 3.**
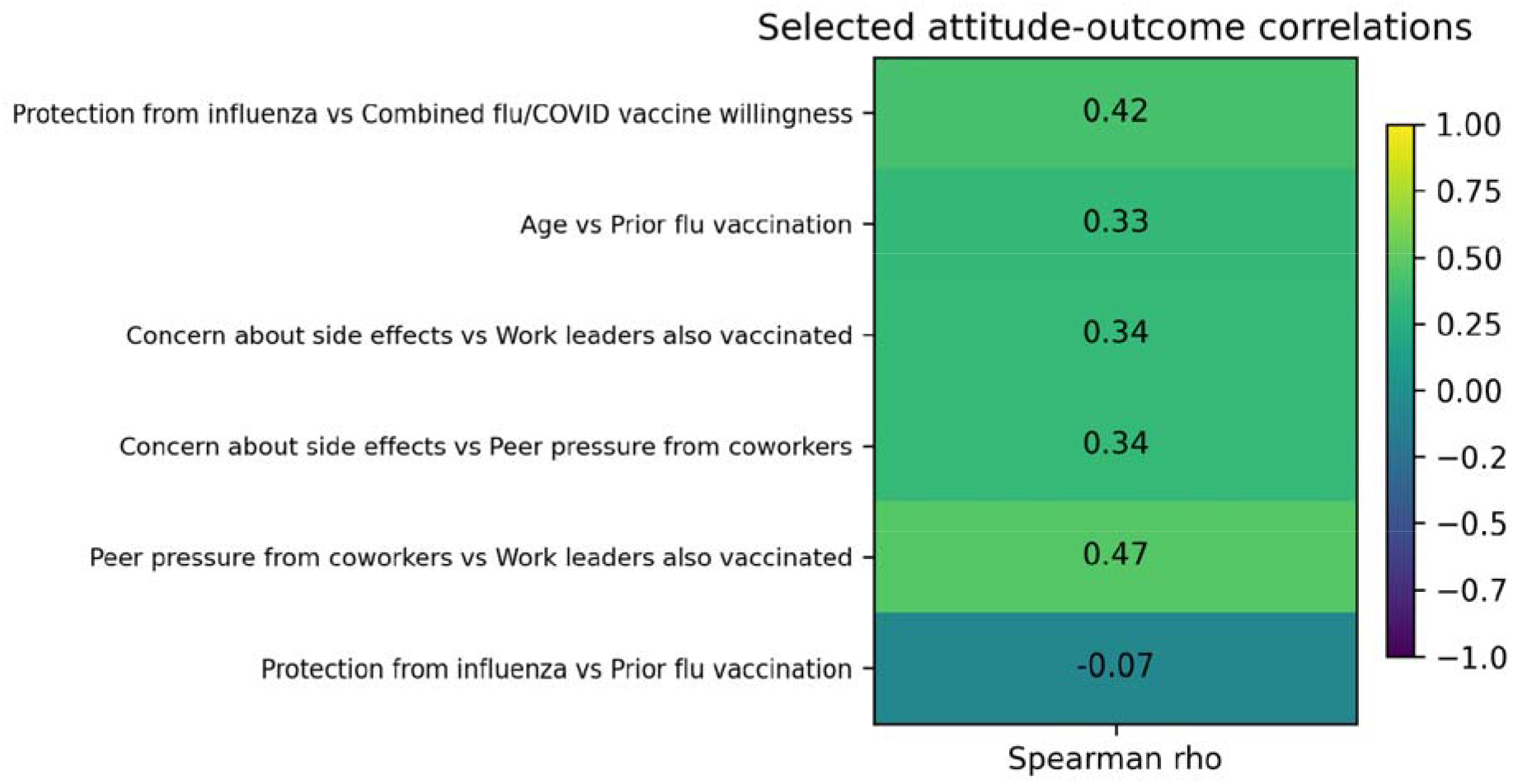
Selected Spearman correlations among vaccination attitudes and outcomes.

### Focus group findings

Focus group findings revealed that the common reasons for employee vaccination included mandatory vaccination requirements, protection of self and others, routine annual vaccination, and avoiding mask requirements. Reasons against vaccination included side effects and lack of trust. Scheduling around work duties was the most frequently reported operational barrier.

## Discussion

This study found that the convenience of on-site vaccination and perceived protection from influenza were the strongest reported drivers of vaccination decisions among healthcare employees. In contrast, peer pressure and small incentives were rated as less influential. These findings suggest that reducing logistical barriers and emphasizing personal health protection may be more effective than relying on social influence in employer-sponsored vaccination programs.

The high rating for convenience is consistent with prior evidence that workplace vaccination campaigns can improve uptake by reducing logistical barriers [2, 3]. Focus-group findings supported this interpretation: employees described scheduling difficulties, limited access during work hours, and the need to leave clinical areas as barriers to vaccination. Conversely, readily available on-site vaccination was viewed favorably.

Exploratory analyses did not identify strong demographic predictors of vaccination attitudes, particularly after accounting for the small sample size and multiple comparisons. Operationally, this suggests that broad access-based interventions may be preferable to strategies that assume hesitancy is concentrated within a specific demographic subgroup.

Overall, the findings support a practical, worker-centered approach to employer-based vaccination programs. Vaccines should be offered close to employee work areas, across multiple shifts, with minimal waiting time and administrative burden. Mobile vaccination teams may be particularly useful for hard-to-reach groups. Education about vaccine safety and benefits remains important, especially given reported concerns about side effects, but it should be paired with convenient access. The central implication is that vaccination programs should be designed around employees’ work schedules and clinical realities rather than around the convenience of the vaccination clinic.

## Limitations

This study was conducted in a single academic healthcare system and used a voluntary survey design, which may limit generalizability. The sample size was modest and some demographic fields had missing values. Several statistical tests should be interpreted as exploratory because the study was not powered for subgroup analyses and Likert outcomes are ordinal. Future work should include larger samples, longitudinal vaccination outcomes, and pre-specified multivariable models.

## Conclusions

In this survey of healthcare employees, convenience and perceived protection were the most important influenza vaccination decisions. Employer-sponsored on-site vaccination programs address logistical barriers and may improve vaccine uptake. Future interventions should pair convenient vaccine access with targeted communication addressing protection, side effects, and trust. Designing vaccination programs around employee workflows—not the workflow of the vaccination clinic—may maximize participation and strengthen employer-based vaccination initiatives.

## Acknowledgements

Affiliation: Center for Occupational and Environmental Medicine, UC San Diego Health, 330 Lewis Street, San Diego, CA 92103.

Grant support: ACOEM Vaccine Initiative is supported by the Centers for Disease Control and Prevention / U.S. Department of Health and Human Services / Council of Medical Specialty Societies. The content is solely the responsibility of the authors and does not necessarily represent the official views of, nor endorsement by, CDC/HHS/CMSS or the U.S. Government.

Dr. Sitapati’s work is supported by the Lawrence S. Friedman Professor of Population Health Endowed Chair.

## Data Availability

The datasets generated and analyzed during this study are not publicly available because they contain confidential employee information collected as part of an internal quality improvement and program evaluation initiative. Access to these data is restricted under institutional privacy policies.

## Ethics Statement

This project was conducted as part of the ACOEM Vaccine Initiative at UC San Diego Health. UC San Diego Health determined that the local activities constituted a quality improvement and program evaluation initiative and did not meet the federal definition of human subjects research under 45 CFR 46; therefore, local Institutional Review Board (IRB) review was not required. The parent ACOEM Vaccine Initiative, supported by the Centers for Disease Control and Prevention (CDC), was conducted under IRB oversight. Survey participation was voluntary and anonymous.

## Contributions

Amy M. Sitapati conceived the project and contributed to the authorship of the manuscript.

## Address

## References

1. Li T, Qi X, Li Q, Tang W, Su K, Jia M, Yang W, Xia Y, Xiong Y, Qi L, Feng L. A systematic review and meta-analysis of seasonal influenza vaccination of health workers. Vaccines (Basel). 2021;9(10):1104. doi:10.3390/vaccines9101104.

2. Gualano MR, Santoro PE, Borrelli I, et al. Employee participation in workplace vaccination campaigns: a systematic review and meta-analysis. Vaccines (Basel). 2022;10(11):1898.

3. Lee BY, Bailey RR, Wiringa AE, et al. Economics of employer-sponsored workplace vaccination to prevent pandemic and seasonal influenza. Vaccine. 2010;28(37):5952–5959.

4. Lee JT, Hu SS, Zhou T, et al. Employer requirements and COVID-19 vaccination and attitudes among healthcare personnel in the U.S.: findings from National Immunization Survey Adult COVID Module, August-September 2021. Vaccine. 2022;40(51):7476–7482.

5. Naleway AL, Henkle EM, Ball S, et al. Barriers and facilitators to influenza vaccination and vaccine coverage in a cohort of health care personnel. Am J Infect Control. 2014;42(4):371–375.

